# Real-Time Monitoring of COVID-19 in Scotland

**DOI:** 10.1101/2020.07.30.20158790

**Authors:** Giles David Calder-Gerver, Stella Mazeri, Samuel Haynes, Camille Anna Simonet, Mark EJ Woolhouse, Helen Brown

## Abstract

To manage the public health risk posed by COVID-19 and assess the impact of interventions, policy makers must be able to closely monitor the epidemic’s trajectory. Here we present a simple methodology based on basic surveillance metrics for monitoring the spread of COVID-19 and its burden on health services in Scotland. We illustrate how this has been used throughout the epidemic in Scotland and explore the underlying biases that have affected its interpretation.

## Responding to the COVID-19 Pandemic

The COVID-19 pandemic has highlighted the importance of timely interventions in responding to health crises. In order to assess the impact of these interventions, methods for monitoring disease spread and burden must be easy to interpret and convey the trajectory of epidemics in real time. We propose a simple approach based on ratios of frequencies over a rolling two-week period as a suitable metric in the context of the Scottish Government’s response to COVID-19.

The first confirmed case of COVID-19 in Scotland was recorded on 1st March 2020 ^[1]^ and Scotland, along with the rest of the UK, entered lockdown on 23rd March 2020 ^[2,3]^. As lockdown measures took effect and the transmission rate of SARS-CoV-2 decreased, we required a method to infer the prevailing trajectory of the epidemic from data that were available from Health Protection Scotland (HPS) on a daily basis ^[1]^.

## Determining a Surveillance Method

Publicly available data on: confirmed cases, patients in hospital, patients in intensive care units (ICUs), and deaths were used to generate our indicators. The cumulative number of COVID-19 cases and deaths have been reported on a daily basis since the start of the epidemic by HPS ^[1]^. Deaths are defined as those which have been registered with the National Records of Scotland (NRS) where a positive test for COVID-19 was recorded in the 28 days prior to death. The NRS additionally reports all COVID-19 deaths (i.e. where COVID- 19 was confirmed or suspected) on a weekly basis ^[4]^ but this was not sufficiently up-to-date to use for surveillance purposes. The number of COVID-19 patients in hospital and in ICUs at midnight is provided by NHS health boards on a daily basis. (See Technical Appendix for further detail).

We sought a reliable method to monitor the epidemic’s trajectory. For both confirmed cases and deaths, we found that methods based on daily counts, e.g. growth rates, were affected by systematic low reporting of counts at weekends. We therefore chose to calculate ratios of the sum of new cases or deaths in the past week compared to the corresponding sum in the previous week. Weekly ratios are a versatile and easily understood method for tracking multiple impacts of COVID-19, and have previously been proposed for the rapid detection of pandemic influenza ^[5]^.

Confidence intervals for the ratios (w1/w2) were calculated based on the approximate standard error for their logs, v{1/w1 + 1/w2}, where w1 = total cases in past week and w2 = total cases in week prior (see Technical Appendix). Whilst weekly ratios were robust to weekday variations, they were still subject to the effects of low reporting on public holidays (when most registries were closed). We made a simple manual adjustment for this by apportioning the number of deaths reported on Mondays and Tuesdays of ‘holiday weeks’ to match the distribution of deaths observed on Mondays and Tuesdays of ‘non-holiday weeks’ (see Technical Appendix).

A similar method was used for patients in hospital and in ICUs. As patients were included in consecutive counts over several days, the ratio for these metrics was based on the count on a given day compared to the count one week before.

## Monitoring COVID-19 in Scotland

The overall trends shown in Figure 1 are similar. All four ratios are clearly above 1 in late March, falling to values of approximately 1 in mid-April and maintaining values of less than 1 from late April onwards, indicating that the epidemic had passed its peak.

**Figure 1:**
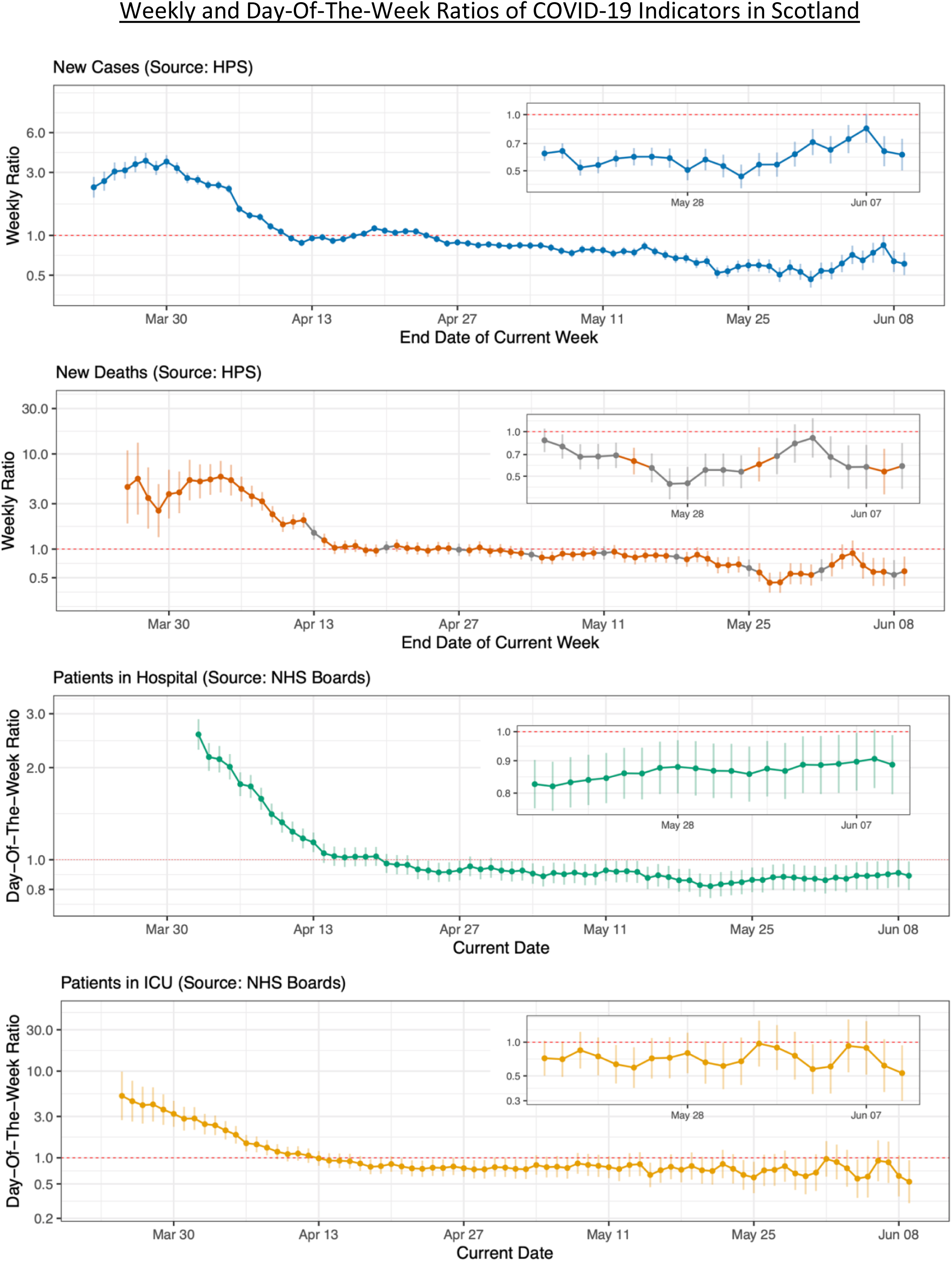
Weekly ratios for new cases and new deaths are ratios of weekly sums, plotted on a log10 scale. Day-of-the-week ratios for patients in hospital and ICU are ratios of daily counts, measured 7 days apart and plotted on a log10 scale. Entries in grey are adjusted for public holiday effects. The series for Patients in Hospital starts later than others as numbers of confirmed cases were not available until March 27^th^. Figure insets highlight ratios and trends for the last 21 days.

The ratio for cases was the first ratio to fall below 1, on 11^th^ April, but did not stay consistently below 1 until after 25^th^ April. This pattern may have been influenced by the increase in testing that occurred during that period.

Ratios for deaths first fell below 1 on 19^th^ April, and stayed at around this level for a further two weeks before becoming consistently lower in mid-May. This delayed decline is likely to have been influenced by the fact that the peak for deaths occurred later in care homes than in other settings (Figure 3), and also by the increasing proportion of ‘tested’ deaths being included in the HPS counts (see Figure 2, where HPS ratios are noticeably higher than those for NRS in late April and early May).

**Figure 2:**
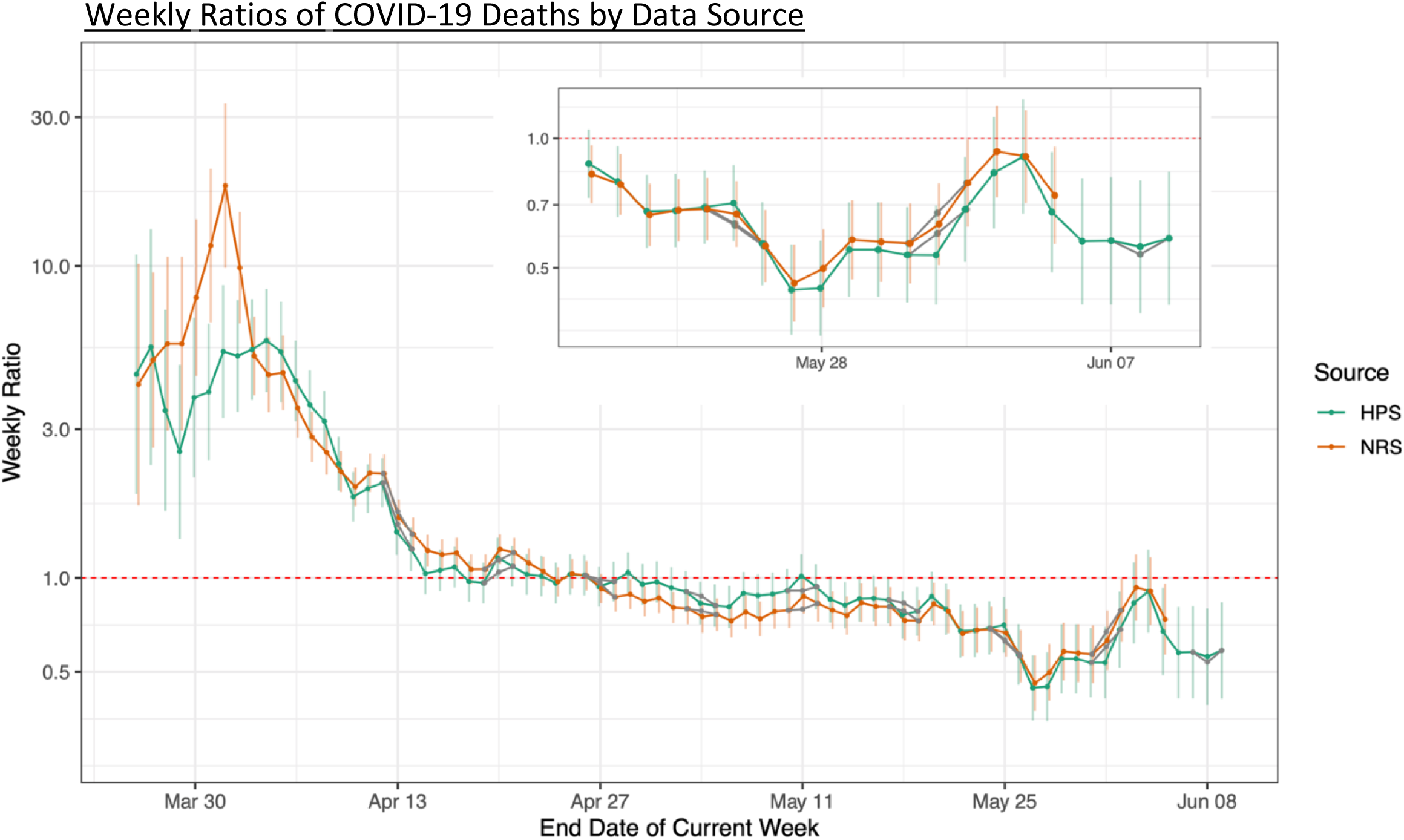
Weekly ratios of new deaths, compared by data source. Entries in grey are adjusted for public holiday effects. Figure inset highlights ratios and trends for the last 21 days.

**Figure 3:**
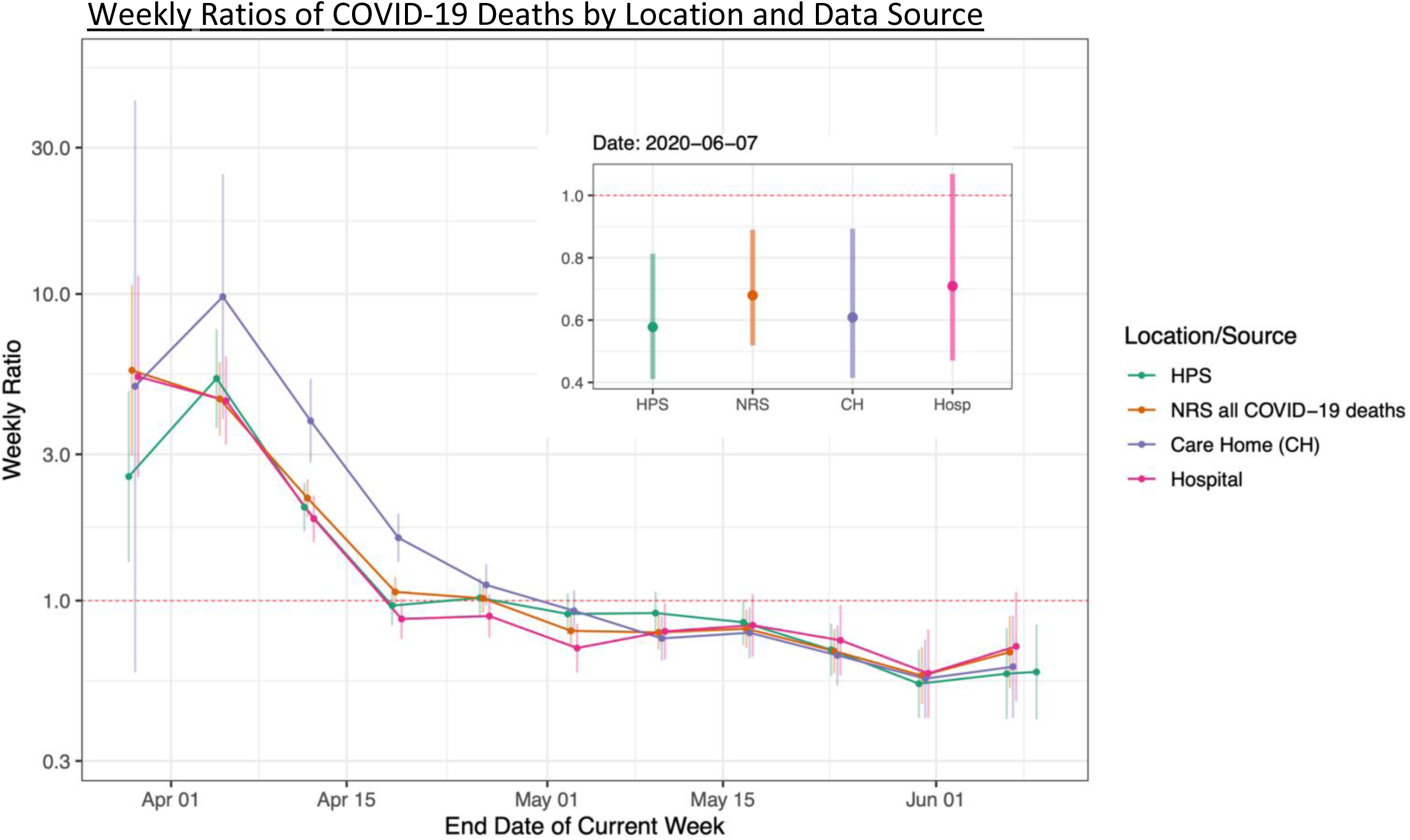
Weekly ratio of new deaths, compared by location and data source. Figure inset highlights the most recent date on which both HPS and NRS data were available. A breakdown of deaths by location (such as care homes and hospitals) is provided in the weekly NRS report. Frequencies for other locations (e.g. home) were too small to provide meaningful ratios.

Ratios for patients in hospital were the last of the four metrics to decline and did not fall below 1 until 20^th^ April. However, this may be partly due to changes in policies for admitting and discharging patients resulting in longer durations of stay (e.g. the requirement for two negative tests for discharging care home residents).

Ratios for patients in ICU first fell below 1 on 14^th^ April and have remained below 1 since. However, this metric predominantly reflects cases occurring in the community rather than in care home residents as very few patients aged over 85 were admitted to ICU.

Despite the inherent biases in the underlying metrics, taken together, the four metrics do indicate that the rate of spread of COVID-19 in Scotland decreased considerably since lockdown was introduced and that, from late April to early June, the epidemic has been in consistent decline. However, there was a notable lag of 3-4 weeks after lockdown before the metrics fell below 1.

The NRS death records facilitate a comparison of COVID-19 deaths in different locations, such as care homes and hospitals (Figure 3). Ratios for care homes had the highest values in early April, and took the longest time to fall below one, highlighting a greater initial rate of spread of SARS-CoV-2 and a later peak for this sector. By contrast, ratios for hospital deaths fell below 1 almost two weeks earlier, indicating that cases in the community peaked earlier than cases in care homes.

These outputs formed part of a report to the Scottish Government’s COVID-19 Advisory Group on a twice-weekly basis. The ratios were straightforward for officials to interpret and contributed to the body of evidence considered when advising on the easing of lockdown ^[6]^.

## Discussion

We have demonstrated that weekly ratios are a versatile and easily understood method for tracking multiple impacts of COVID-19. However, as can be seen in the four metrics considered here, different data sources are susceptible to different biases and these must be clearly communicated to policy makers. For cases and deaths, changes in testing policy and coverage can affect the like-for-like comparability of these metrics over time. If test coverage is increased then an increased proportion of clinically milder and previously overlooked cases is likely to be detected. Deaths counted by HPS are also directly affected by increased testing. Metrics may also be influenced by changes to policy, e.g. changes to policy regarding the hospitalisation, testing and discharge of elderly people and care home residents were implemented in late April ^[7,8]^.

As deaths are generally less affected by changes in testing policy than cases, it could be argued that death ratios may be the most consistent indicator of COVID-19, and possibly SARS-CoV-2, spread. However, the number of new deaths is heavily influenced by the rate of spread to and within groups who are predisposed to negative clinical outcomes, such as the elderly and those with underlying medical conditions, many of whom live in communal settings and are dependent on close physical care. Ratios for deaths will therefore intrinsically reflect the rate of spread in these higher risk groups more than in lower risk groups. Other measures are similarly weighted towards more severely ill patients: cases only include people who have been able to get tested, and patients in hospital and ICU will only include those with sufficiently severe symptoms to require admission.

The R value has become an important metric in social and political settings to express the rate of spread of COVID-19. The objective to keep it below 1 is straightforward to understand. However, its current value is challenging to estimate accurately from available statistics ^[9]^. The metrics described here are much more straightforward to calculate and, when interpreted appropriately, can be used to assess the efficacy of lockdown measures and inform decisions about the easing of lockdown restrictions. They could easily be adapted for use in other countries reporting similar data.

## Data Availability

All data we use are publicly available, from the National Records of Scotland or Health Protection Scotland (via The Scottish Government). During the first ∼3 weeks of data collection (before time series data were available) this data was collated by web scraping. Scraped data was also used to cross-check official running totals.

https://github.com/gcalder/COVID-19

https://www.nrscotland.gov.uk/covid19stats

https://www.gov.scot/publications/coronavirus-covid-19-daily-data-for-scotland/

https://www.gov.scot/publications/coronavirus-covid-19-trends-in-daily-data/

## Conflicts of Interest

None declared.

## Funding

This work was supported by a Wellcome Trust ISSF award through the University of Edinburgh.

## Data Availability

All data analysed by this study are publicly available from The National Records of Scotland or Health Protection Scotland, via The Scottish Government.

## Technical Appendix

The code used to produce our twice-weekly report, and a similar report we distribute on a daily basis, is available at https://github.com/gcalder/COVID-19.

### Data Sources

The cumulative number of COVID-19 cases and deaths has been reported since the start of the epidemic by HPS on a daily basis. Deaths are defined as those which have been registered with the National Records of Scotland (NRS) where a positive test for COVID-19 was recorded in the 28 days prior to death. These numbers are expected to capture the majority of COVID-19 deaths occurring in hospitals, but a lower proportion of deaths outwith hospital settings, especially in late March and early April (when testing was mostly restricted to hospitalised patients ^[10,11]^). The NRS additionally report all COVID-19 deaths mentioned in the death certificate (i.e. where COVID-19 was confirmed or suspected) and therefore capture a greater proportion of deaths in care homes and the wider community. However, this report was only issued on a weekly basis and was not sufficiently up-to-date to use for surveillance purposes. The number of COVID-19 patients in hospital and in ICUs at midnight is reported by NHS health boards on a daily basis. We included both confirmed and suspected patients in the ICU numbers, but, due to reporting inconsistencies, only included confirmed patients in the patients in hospital figures.

### Confidence Interval Calculation

Confidence intervals for the ratio have been calculated based on the standard error of the log of the ratio (w1/w2) which is approximated by {1/w1 + 1/w2}, based on assuming a Poisson distribution for w1 and w2 (where w1 = total cases in past week, w2 = total cases in previous week). The approximation is obtained from the standard formula:

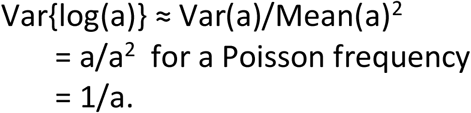

The ratio on a log scale may be expressed:

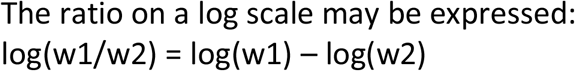

giving

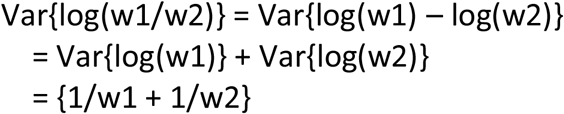

and

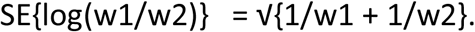

The 95% confidence interval is then calculated as log(w1/w2) ± 1.96 x SE, which is exponentiated to provide the confidence intervals displayed.

### Public Holiday Adjustment

Most registries are closed on public holidays so deaths cannot be registered. This causes a bias in the resulting ratio for deaths. For example, a low number of deaths were registered on 4th May (Spring bank holiday), followed by a high number on Tuesday 5th May. This caused the ratio for Monday 4th May to be artificially low, the ratio the following Monday (11th May) to be artificially high, and the ratio on the Monday two weeks later (18th May) to be artificially low. This was due to the separation of the (low and high) numbers for Monday 4th and Tuesday 5th May between the numerator and denominator for ratios on these Mondays. A similar pattern was noticed for other public holidays, e.g. 13th April (Easter Monday) and 18th May (Victoria Day), which affected ratios on the holiday dates and on the two subsequent Mondays in the same way. We made a simple manual adjustment for this bias by apportioning the number of deaths reported on Mondays and Tuesdays of ‘holiday’ weeks to match the distribution of those observed on Mondays and Tuesdays of ‘non-holiday’ weeks. For example, 44 and 83 deaths were reported by HPS for Monday 4th and Tuesday 5th May and these 127 deaths were redistributed to be 60 and 67, so that they followed the average proportions observed for ‘non-holiday’ Mondays and Tuesdays (47.1% and 52.9%).

## Notes

### Competing Interest Statement

The authors have declared no competing interest.

